# Long-term neutralizing antibody dynamics against SARS-CoV-2 in symptomatic and asymptomatic infections: a systematic review and meta-analysis

**DOI:** 10.1101/2022.12.15.22283503

**Authors:** Wanying Lu, Nan Zheng, Xinhua Chen, Ruijia Sun, Jiayi Dong, Shijia Ge, Xiaowei Deng, Hongjie Yu

## Abstract

**Background:** The kinetics of the neutralizing antibody response against SARS-CoV-2 is crucial for responding to the pandemic as well as developing vaccination strategies. We aimed to fit the antibody curves in symptomatic and asymptomatic individuals.

**Methods:** We systematically searched PubMed, Embase, Web of Science, and Europe PMC for articles published in English between Jan 1, 2020, and Oct 2, 2022. Studies evaluating neutralizing antibody from people who had a natural SARS-CoV-2 infection history were included. Study quality was assessed using a modified standardized scoring system. We fitted dynamic patterns of neutralizing antibody using a generalized additive model and a generalized additive mixed model. We also used linear regression model to conduct both univariate and multivariable analyses to explore the potential affecting factors on antibody levels. This study is registered with PROSPERO, CRD42022348636.

**Results:** 7,343 studies were identified in the initial search, 50 were assessed for eligibility after removal of duplicates as well as inappropriate titles, abstracts and full-text review, and 48 studies (2,726 individuals, 5,670 samples) were included in the meta-analysis after quality assessment. The neutralization titer of people who infected with SARS-CoV-2 prototype strain peaked around 27 days (217.4, 95%CI: 187.0-252.9) but remained below the Omicron BA.5 protection threshold all the time after illness onset or confirmation. Furthermore, neither symptomatic infections nor asymptomatic infections could provide over 50% protection against Omicron BA.5 sub-lineage. It also showed that the clinical severity and the type of laboratory assays may significantly correlated with the level of neutralizing antibody.

**Conclusions:** This study provides a comprehensive mapping of the dynamic of neutralizing antibody against SARS-CoV-2 prototype strain induced by natural infection and compared the dynamic patterns between prototype and variant strains. It suggests that the protection probability provided by natural infection is limited. Therefore, timely vaccination is necessary for both previously infected symptomatic and asymptomatic individuals.

## Introduction

Corona Virus Disease 2019 (COVID-19) is an acute respiratory infectious diseases caused by severe acute respiratory syndrome coronavirus 2 (SARS-CoV-2). Parts of COVID-19 patients develop symptoms such as fever, cough, and headache, but more individuals may show no symptoms all the time. These asymptomatic individuals account for a larger proportion yet they are harder to be found^1^. From the first outbreak in 2020 to the present, over 615 million cases have been caused^2^ and the pathogen, has undergone several mutations and derives many variants over time as well. WHO has paid special attention to Alpha (B.1.1.7), Beta (B.1.351), Gamma (P.1), Delta (B.1.617.2) and Omicron (B.1.1.529), which has been listed as Variants of Concern (VOCs)^3^. In January 2022, Omicron BA.1 sub-lineage was taking over the Delta variant and becoming dominant globally. Gradually, BA.2 and its constituent sub-lineages, such as BA.2.12.1, are overtaking BA.1 as the dominant variant globally. More recent two sub-lineages BA.4 and BA.5 gradually became prevalent since June 2022, with the BA.5 being the dominantly circulating strains around the world.

Existing evidence suggests that there are more than 60 substitutions/deletions/insertions in Omicron^4^, with fifteen mutations located in the receptor binding domain (RBD), a region that mediates virus attachment to the angiotensin converting enzyme 2 (ACE2) receptor on target cells and is the principal target of neutralizing antibody that contributes to protection against SARS-CoV-2^5,6^. These mutations give Omicron variant increased transmissibility and enhance its immune-escape ability, which made it the major circulating variantcurrently^3^.

After infected with SARS-CoV-2, individuals produce neutralizing antibody, which is part of the humoral response and prevents the interaction of infectious particles with host cells by interfering with virion binding to receptors and blocking virus uptake into host cells^7^. Therefore, neutralizing antibody produced in infected individuals may protect them from reinfection to a certain extent. In other words, neutralizing antibody is likely to be a key correlate of immune protection^8^. However, neutralizing antibody level may be affected by age and clinical severity, and as neutralizing antibody titer declines over time, its protection diminishes as well^9^. When neutralizing antibody drop below a certain level, reinfection may occur^10^. To maintain effective neutralizing antibody levels, vaccination is a good way. For the general population, a booster dose or even a fourth dose is recommended nowadays. However, for individuals who have ever been infected, only a few countries have defined clear vaccination strategies and these strategies differ a lot across settings. For example, China recommends COVID-19 convalescents to vaccinate one dose six months after infection^11^, while the United States recommends delaying the next dose for three months after infection^12^.

Considering the limited vaccine resources and the fairness of distribution, when and how to vaccinate are urgent issues for COVID-19 convalescents, which requires studies on antibody kinetics and immune protection in natural SARS-CoV-2 infections. At present, there are few reviews that quantitatively and comprehensively summarized the long-term dynamics of neutralizing antibody titer in COVID-19 convalescents or asymptomatic infections, neither for prototype strain nor for other variants. The existing systematic reviews and meta-analysis are based on limited numbers of studies, which make the summary of the antibody kinetics limited to the first few months after infection and lack comparability between different virus neutralization assays^13–16^.

In this study, we systematically collected data on time-varying neutralizing antibody from people who had a natural SARS-CoV-2 infection history, including symptomatic COVID-19 convalescents and asymptomatic infections. Based on collected data, we fitted dynamic patterns of neutralizing antibody using a generalized additive model (GAM) and a generalized additive mixed model (GAMM), estimated the peak times and detectable duration and explored the potential effects that age group, clinical severity and neutralization assays on antibody levels using both univariate and multivariable analyses. We also compared the differences showed in kinetic patterns between the prototype strain and variants.

## Methods

### Search strategy and selection criteria

According to the Preferred Reporting Items for Systematic Reviews and Meta-Analyses protocols^17^, we systematically did a literature review from three peer-reviewed databases (PubMed, Embase, and Web of Science) and one preprint sever (Europe PMC). The key search terms used were as follows: “SARS-CoV-2”, “neutralizing antibody”, “dynamic”, “kinetics”, “symptomatic”, and “asymptomatic”. Detailed search terms were listed in **Supplementary Table 1**. Two independent researchers (WL, NZ) assessed all retrieved papers published in English from Jan 1, 2020 to Oct 2, 2022. A third researcher (XC) was consulted when the two reviewers disagreed on study assessment. We included studies that reported neutralizing antibody against SARS-CoV-2 prototype or variant strains by using serum or plasma collected from symptomatic and asymptomatic individuals with virologically or serologically-confirmed SARS-CoV-2 infections. Studies that investigated immunogenicity among specific population, such as pregnant women, cancer people, immunodeficient patients, would be excluded. Detailed inclusion and exclusion criteria were listed in **Supplementary Table 2**. This study was registered with PROSPERO (no. CRD42022348636).

### Quality assessment

Based on the Newcastle–Ottawa Scale (NOS)^**18**^ scoring system recommended by the Cochrane Collaboration, we developed a revised scoring system to assess the study quality. We comprehensively assessed study design (e.g., clear criteria and definition), laboratory assay (e.g., validated neutralization assay), and outcome interpretation (e.g., clear definition of sampling time) for each included study. The scoring system has a total of nine items, with one point assigned for each item. A score of 0–3 was considered a low-quality (grade C) study, a score of 4–6 was considered a moderate-quality (grade B) study, and a score of 7–9 was considered a high-quality (grade A) study Detailed scoring system were listed in **Supplementary Table 3**. Only high and moderate-quality literatures were included in this study.

### Data extraction

For eligible studies, two researchers (WL, NZ) extracted the following data using a standardized electronic data collection form: published journal or preprints severs, title, first author’s name, study location, study design, study participants, sample sizes, sampling time, age, sex, race, ethnicity, underlying conditions, clinical severity, details of laboratory testing methods including neutralization assays used, sample type (sera or plasma), and other factors that may affect the serological results, such as dilution factor, number of duplicates, cell line, virus titer used in the assay and the individual antibody titer that neutralizes or inhibits 50% of the virus(e.g., NT_50_, PRNT_50_). For each included study, we summarized its study participants, laboratory methods, and outcome in **Supplementary Table 4**. Pre-trained researchers digitized individual titer values from the figures in articles if titer were not available in table format^19^. We made reliable assumptions according to the information of the locally circulating viruses and the time when the variants begin to circulate in the region if the exact virus lineage causing the infection for some participants is unknown. Besides, if key information, such as the details of laboratory testing method, was not reported in the paper, we will contact the corresponding author via email for data inquiry. For those individuals with titer lower than limit of detection, we assumed a titer half that limit (e.g., a titer of 10 is assumed when “<20” was present)^20^. For those individuals with titer higher than limit of detection, we assumed a titer that limit (e.g., a titer of 1024 is assumed when “>1024” was present).

### Data classification and synthesis

We classified multiple neutralization assays used in included studies into three categories based on the type of virus (live or pseudo) used and the types of vectors (lentivirus or vesicular stomatitis virus [VSV]) used in pseudovirus neutralization assays: live virus neutralization assays, lentivirus-vector pseudovirus neutralization assays, and VSV-vector pseudovirus neutralization assays. In each category of assay, we further stratified study participants into two groups based on the presence or absence of symptoms: (1) symptomatic individual^21^: a symptomatic individual is one with laboratory-confirmed SARS-CoV-2 infection as determined by PCR and/or serology and with symptoms including but not limited to fever, cough, general weakness/fatigue, headache, myalgia, sore throat, coryza, dyspnea, anorexia/nausea/vomiting, diarrhea. (2) asymptomatic individual^22^: an asymptomatic individual is one with laboratory-confirmed SARS-CoV-2 infection as determined by PCR and/or serology but with no symptoms whatsoever for the duration of infection. The characteristics of the participants (e.g., age, clinical severity and comorbidities) were defined as follows. We divided the ages into three groups: ≤15, 16-60 and >60. As for clinical severity in symptomatic cases, we distinguished between mild and severe individuals according to the World Health Organization^23^ and National Institutes of Health^24^. Standards were as follows: (1) severe case: the patient were categorized into severe cases if they were attended to Intensive Care Unit (ICU), received oxygen supplementation (decreased oxygen saturation (≤94%)), ventilation treatment during the hospitalization, or developing of pneumonia. (2) mild case: symptomatic but not severe case was classified as mild case. For comorbidities, we only distinguished whether they have comorbidities instead of what or how many comorbidities there were.

### Data analysis

Titer of symptomatic and asymptomatic infection-induced SARS-CoV-2 neutralizing antibody were log-transformed before data analysis. Model fitting was carried out according to the types of virus strains, which confirmed by article report or gene sequencing. A generalized additive model with Gaussian distribution that allows for flexible specification of dependence of response to covariates was used to fit the kinetics of SARS-CoV-2 neutralizing antibody. The model was shown in Equation 1:

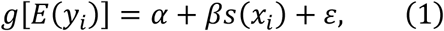

Where *E*(*y*_*i*_) is mean neutralizing antibody titer against SARS-CoV-2, *g* is the log link function, *α* is the intersect, *s*(*x*_*i*_) is the value at a given time *x*_*i*_ of a smoothing spline basis, *β* is the vector of spline coefficient, and *ε* is the random error.

Considering of the individual random effects, we also presented a subset of matched data of which sera were consecutively collected from the same individual, using generalized additive mixed model to model the dynamic curves. We assumed the Gaussian distribution of neutralizing antibody titer in symptomatic individuals while Gamma distribution in asymptomatic individuals based on the type of distribution. The specification for the GAMM smoothing model is defined as follows:

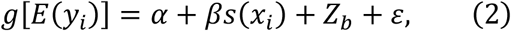

Where *Z*_*b*_ is the random effect. Comparisons between models were made based on Bayesian information criteria (BIC) and Akaike information criteria (AIC). Besides, we determined prototype strain and Omicron BA.5 sub-lineage protective threshold which defined as 50% protective neutralization level against virus infections based on previous studies^25,26^.

We used linear regression model to conduct both univariate and multivariable analyses. The factors included age, sex, comorbidity, clinical severity and assays. For all statistical tests, a two tailed p value of less than 0.05 was considered statistically significant. We also compared the neutralizing antibody dynamic curves among different virus strains based on the same laboratory assay and clinical severity. All statistical analyses were done using R software (version 4.2.0).

## Results

### Study selection and data extraction

We identified a total of 4,550 studies after systematically searching multiple data sources with 1,966 coming from peer-reviewed databases, and 2,584 from preprint server **(Figure 1)**. After screening titles, abstracts, and full-texts, 50 studies containing a total of 2,726 individuals and 5,783 samples were included. After quality assessment, there were 18 grade A (18/50, 36%), 30 grade B (30/50, 60%) and 2 grade C studies (2/50, 4%), and studies classified as grade A and B were included into analysis **(Figure S1, Table S5)**. Among 48 studies, with prototype strain infections the subjects accounted for the most (47 studies; 5,546 of 5,670 samples, 97.8%), followed by Alpha strain infected individuals (2 studies; 110 of 5,670 samples, 1.9%) and Omicron BA.2 sub-lineage infections (1 study; 14 of 5,670 samples, 0.2%). Live virus neutralization assays were most common (27 of 48 studies, 56.3%), followed by lentivirus-vector pseudovirus neutralization assays (15 of 48 studies, 31.2%) and VSV-vector pseudovirus neutralization assays (6 of 48 studies, 12.5%). Most participants were symptomatic cases (46 studies; 2,480/2,690 individuals, 92.2%; 5,281/5,670 samples, 93.1%), while asymptomatic individuals accounted for a relatively small proportion (12 studies; 210/2,690 individuals, 7.8%; 389/5,670 samples, 6.9%).

**Figure 1.**
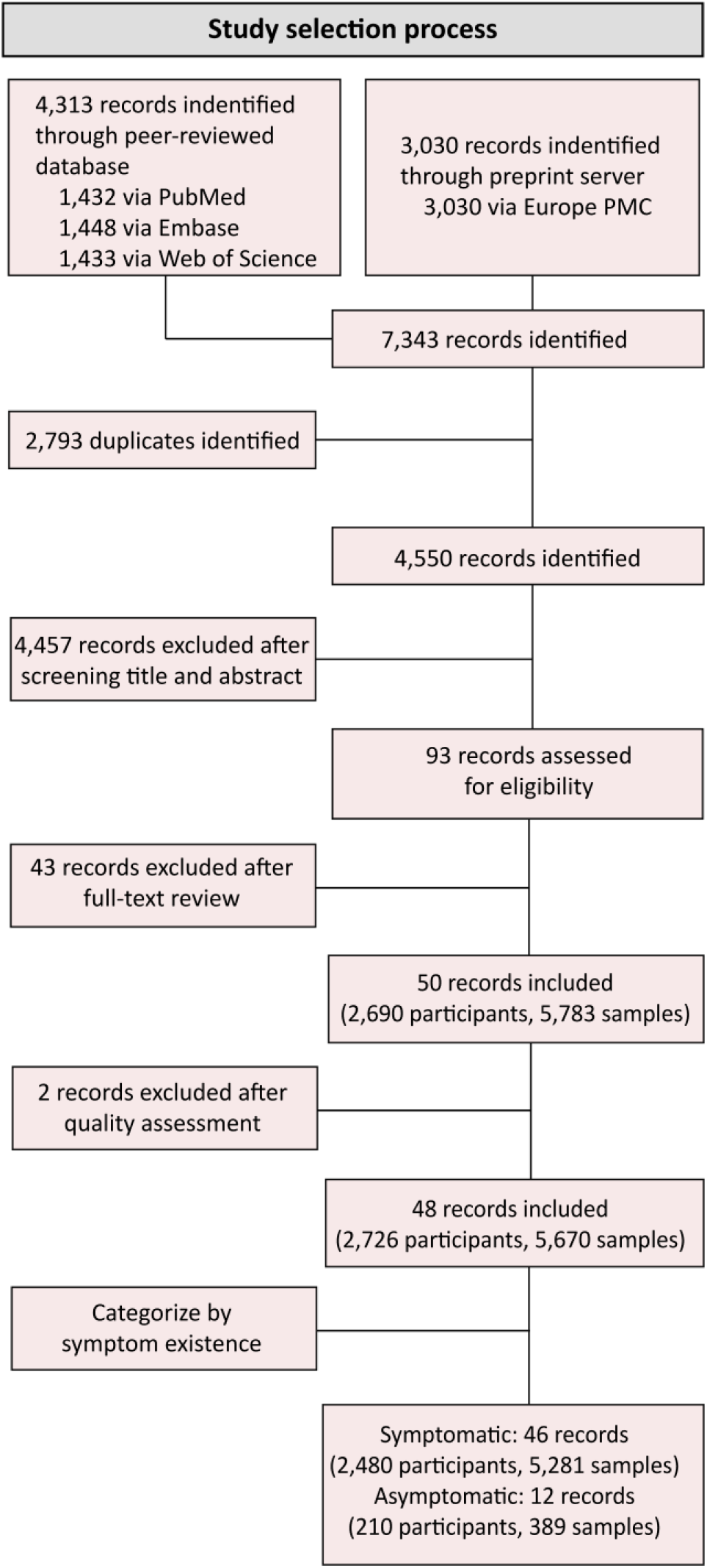
Flowchart of study selection. Flowchart of the selection of studies including time-varying neutralizing antibody data from Jan 1, 2020, to Oct 2, 2022.

### Antibody responses in prototype strain infections in live virus neutralization assays

For the overall dynamic curve fitted with matched sera, titer peaked around 27 days (217.4, 95%CI: 187.0-252.9) after illness onset or confirmation. It remained above the detection limit of 10 after 479 days, dropped below the prototype strain protection threshold (19) at day 467 after illness onset or confirmation and kept below the Omicron BA.5 protection threshold (266) all the time **(Figure 2A)**. As for the dynamic curve in symptomatic and asymptomatic infections separately, antibody titer of symptomatic convalescents peaked around 26 days (250.8, 95%CI: 214.8-292.7) after illness onset, then declined **(Figure 2B)**. Antibody titer of asymptomatic individuals were consistently lower than that of symptomatic convalescents all the time. It took 463 and 171 days for neutralization titer to drop to the prototype strain protection threshold (19) respectively in symptomatic individuals and asymptomatic individuals but both of them remained below the Omicron BA.5 protection threshold (266) and above the detection limit of 10 during the whole follow-up period **(Figure 2C)**. Results of all samples showed similarity to those of paired sera, with slight difference mainly in the timing of peak and protection thresholds **(Figure S2)**.

**Figure 2.**
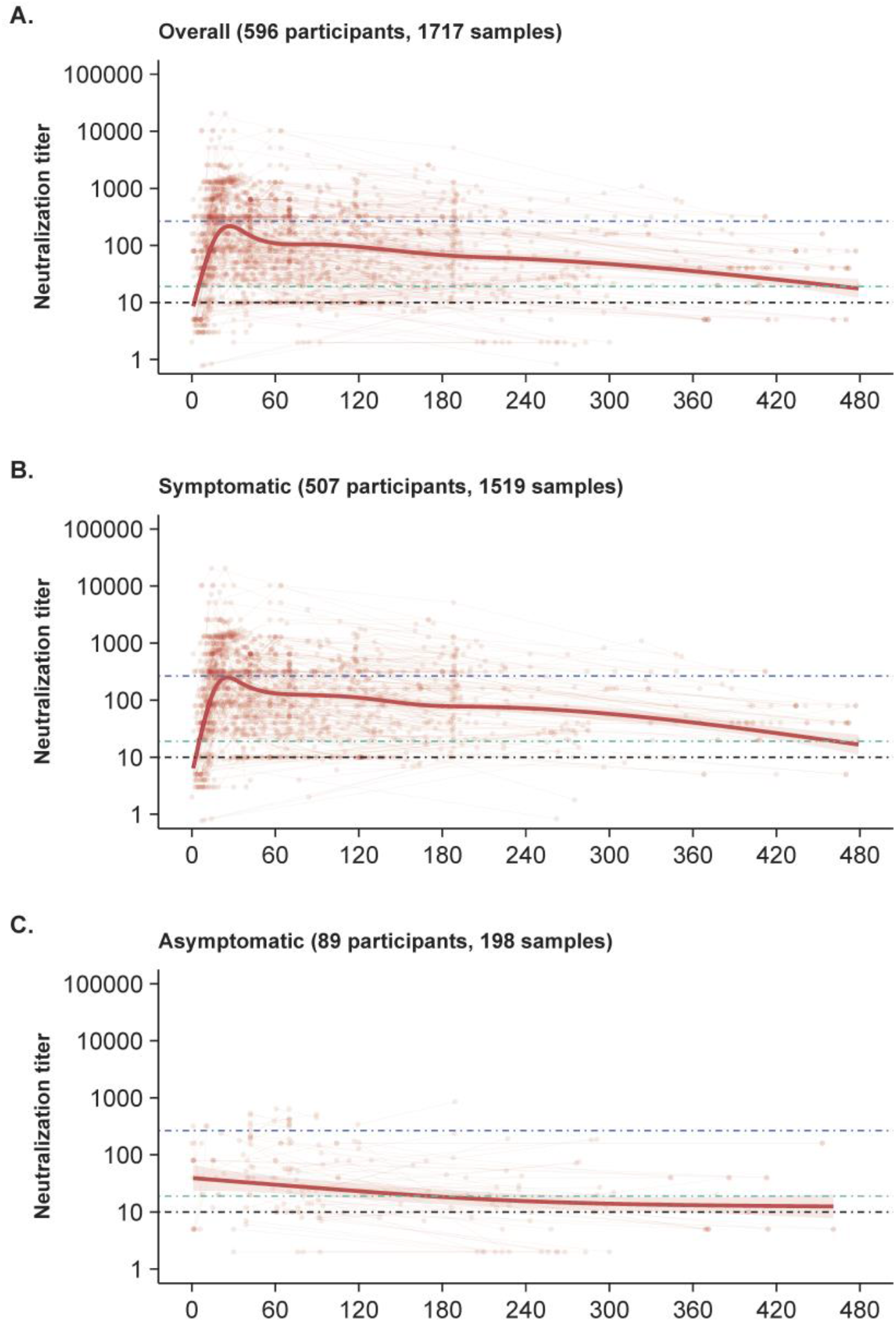
Neutralizing antibody responses in prototype strain infections in live virus neutralization assays. A) All individuals; B) Symptomatic individuals; C) Asymptomatic individuals. Black lines represent detection limit (10); green lines, prototype strain protection threshold (19); blue lines, Omicron BA.5 protection threshold (266).

### Association between clinical outcomes and antibody responses in prototype strain infections

In the univariate analyses, the levels of neutralizing antibody increased over age, with the mean neutralization level of 47.9, 137.6 and 209.2 in the groups of ≤ 15 years, 15-60 years and > 60 years, respectively (**Figure 3, Table S6)**. For different clinical endpoints, the neutralization titer in mild cases was significantly higher than that of asymptomatic individuals, but lower than that of severe cases (p<0.001). The levels of neutralizing antibody varied in the types of neutralization assays, with the neutralizing antibody tested by lentivirus-vector pseudovirus neutralization assays had the highest titer, followed by VSV-vector pseudovirus and live virus neutralization assays. In the multivariable analysis, after controlling potential biases, only clinical severity and types of neutralization assays were statistically significant.

**Figure 3.**
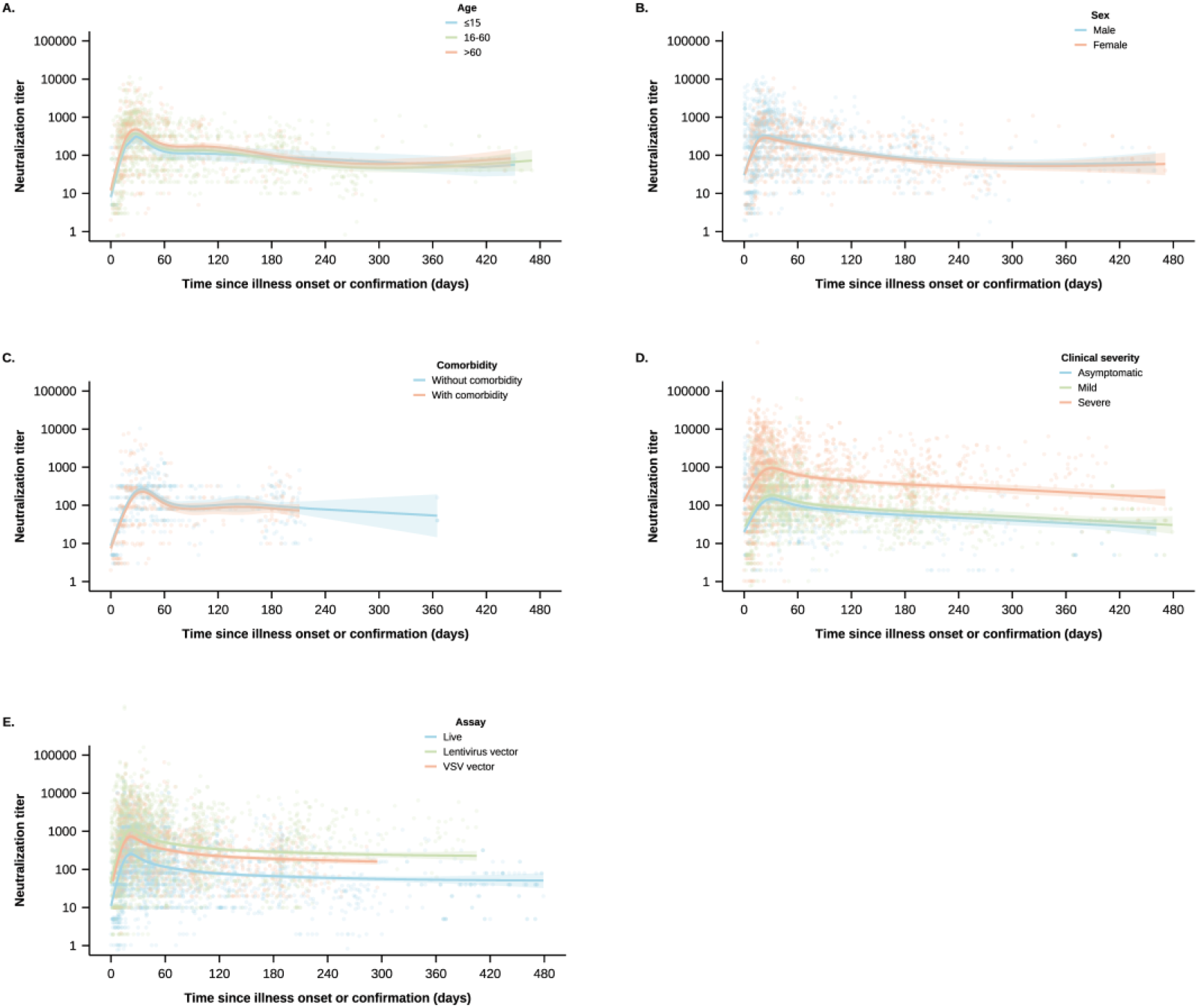
Neutralizing antibody responses in prototype strain infections among different characteristics. A) Age; B) Sex; C) Comorbidity; D) Clinical severity; E) Assay.

### Comparison of neutralizing antibody responses between prototype strain and variants

There are few eligible studies explore the antibody kinetics in individuals infected with variants, with only two studies for Alpha and one study for Omicron BA.2. Considering the limited number of sample size, heterogeneity of study design, as well as different antibody detection method, we compare the pattern rather than the antibody level between prototype strain and variants. As for the comparison between prototype and Alpha strains infections in lentivirus-vector pseudovirus neutralization assays, individual antibody titer was fitted up to 180 days and 122 days for prototype strain and Alpha strain, respectively. Overall, although the follow-up period was shorter in the study of Alpha variant, the dynamic patterns of the both were similar based on matched samples. Antibody titer of Alpha convalescents peaked around 33 days after symptom onset, while peaked around day 28 in prototype convalescents. It kept waning for the next 90 days without signs of plateauing. Up to day 122 after symptom onset, the antibody level of Alpha strain remained higher than that of the prototype strain **(Figure 4)**. As for the comparison between prototype strain and Omicron BA.2 sub-lineage infections in live virus neutralization assays, matched individual antibody titer was available for up to 209 days and 52 days for prototype strain and Omicron BA.2 sub-lineage, respectively. Antibody titer of Omicron BA.2 peaked at day 39, 10 days later than that of prototype convalescents, then showed a downward trend Due to the short follow-up period of the Omicron BA.2 study, it was difficult to compare the complete dynamic patterns with the prototype strain.

**Figure 4.**
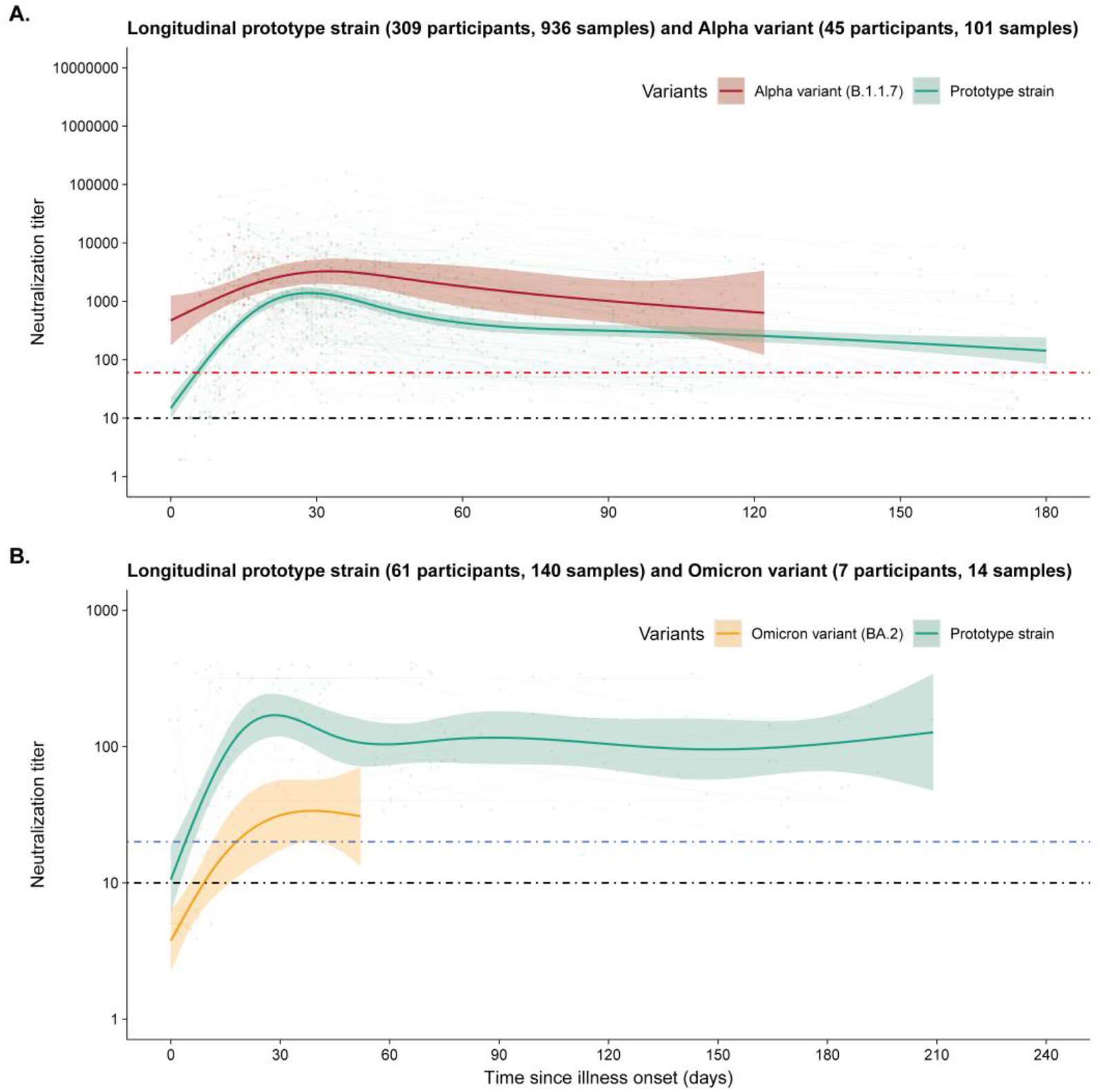
Comparison of neutralizing antibody responses between prototype strain and variants. A) Prototype and Alpha strains; B) Prototype strain and Omicron BA.2 sub-lineage. Black lines represent detection limit of prototype strain (10); red line, detection limit of Alpha strain (60); blue line, detection limit of Omicron BA.2 sub-lineage (20).

## Discussion

Our study systematically retrieved the data of neutralizing antibody titers in naturally infected individuals, fitted the dynamic curves, and explored the factors associated with antibody levels. We also compared antibody responses between prototype and variant strains. We found neutralization titer peaked around 27 days (217.4, 95%CI: 187.0-252.9) after illness onset or confirmation but remained below the Omicron BA.5 protection threshold all the time after illness onset or confirmation. Furthermore, neither symptomatic infections nor asymptomatic infections could provide over 50% protection against Omicron BA.5 sub-lineage. Our results also showed that the clinical severity and the type of laboratory assays may significantly correlated with the level of neutralizing antibody.

Our study showed that neutralizing antibody in natural infection individuals could be detected at around 2 days after confirmation or onset of symptoms, peaked around 27 days, and then started to decline. The antibody level remained detectable (>1:10) for at least 16 months, which was consistent with previous studies that showed that neutralizing antibody could be detected at around 6-15 days after infection^27^, reached peak around 31-45 days^28^, then started to decline^14,16^, and remained near the detection limit at around 360 days^29^. As for the asymptomatic infections, we did not observe a marked increase of antibody level during the acute phase. Since asymptomatic individuals remain asymptomatic from exposure to infection and virus expulsion, these individuals are often difficult to be captured by symptom-based surveillance. In addition, because individuals are asymptomatic all the time, they usually do not initiate to seek assistance in medical institutions, which increases the difficulty of early identification of asymptomatic individuals. Most of the studies included in this study were hospital-based follow-up studies, so it was very hard to identify asymptomatic individuals in the early stage of exposure, nor to better quantify the trend of rapidly increasing neutralizing antibody levels during this period. As for the dynamic patterns of variant strains, we did not compare the antibody levels considering the heterogeneity between studies, such as characteristic of participants and laboratory assays. We only compared the dynamic curves, which were similar to the patterns of prototype strain. Long-term follow-up studies are needed to determine the dynamic curves of the variant strains.

In our study, we found neutralization titer remained under the Omicron BA.5 protection threshold all the time after confirmation. A test-negative, case–control study showed that the protection probability against Omicron BA.5 infection and symptomatic infection was 33.5% (95% CI: 29.3-37.5%) and 38.1% (95% CI: 27.7-46.9%) separately at 493 and 496 days after infection in individuals who previously infected pre-omicron strain^30^. Another study, which estimated the protection effect in a population with hybrid immunity (previous infection and vaccination), showed that the protection effect against BA.5 infection was 51.6% (95% CI: 50.6-52.6%) in Wuhan-Hu-1 strain infected individuals^31^. Results of these studies suggested that the antibody level after natural infection with the prototype strain could not provide 50% protection probability against Omicron BA.5 infection, which were similar to that of our study. So the COVID-19 vaccines should be administered as soon as possible to obtain effective protection. Like a special “infection”, the COVID-19 vaccine effectiveness or efficacy against the prototype strain are often compared with the protection probability of natural infection. One study showed that the effectiveness against BA.5 hospitalization was 47.4% (19.9-65.5%) within 3-4 months and 19.3% (6.3-30.5%) over 9 months^32^. The effectiveness against BA.5 infection would be lower^25^. It was similar to our study, suggesting that neutralizing antibody levels in naturally infected individuals as well as primary vaccinated population may not provide 50% protection probability against BA.5. This was consistent with previous study that showed the probability of protection after natural infection was similar to the effectiveness of two doses vaccination^33^.

Our study showed that neutralizing antibody levels are positively correlated with clinical severity, which was consistent with previous study^34^ Since inflammatory cytokines may be related to higher levels of neutralizing antibody in severe cases. Severe cases tend to have high systemic level of pro-inflammatory cytokines, which has been shown to correlate with antibody levels to COVID-19^35^. Moreover, inflammatory cytokines are closely related to humoral immunity, and studies have shown that IFN-γ^36^, IL-12^37^, and IL-17^38^ play important roles in B-cell development. Age was statistically significant in univariate analysis, indicating older age was associated with higher neutralizing antibody levels, but not in multivariate analysis. This could be because there were more severe cases in the elderly population, and clinical severity is positively correlated with neutralizing antibody levels, so antibody levels are higher in the elderly population.

Our study had several limitations. First, there were heterogeneity among the studies, but we minimized them by conducting subgroup analyses according to the population characteristic and laboratory assays. Second, the exact virus lineage causing the infection for some participants is unknown. However, we have made reliable assumptions according to the information of the local circulating viruses and the time when the variants began to circulate in the region. Third, relatively fewer sera beyond 200 days after onset of illness was collected to define the late antibody waning kinetics. We also have relatively fewer numbers of asymptomatic infections compared to those with symptomatic disease. Thus, our findings on asymptomatic infections need to be interpreted with caution. Studies on the long-term dynamic changes of antibody in asymptomatic infected patients should be strengthened in the future.

In conclusion, our study provided a comprehensive mapping of the dynamic of neutralizing antibody against SARS-CoV-2 prototype strain induced by natural infection and compared the dynamics between protype and variant strain. Our findings suggested that the antibody level of symptomatic cases reached peak level at day 26 and then decreased, but it failed to provide 50% protection probability against Omicron BA.5, same as that of asymptomatic individuals. At present, the key question is to study the long-term dynamic changes of antibody after natural infection of mutant strains so as to understand when and how to vaccinate COVID-19 convalescents, whether widespread immunization would lead to a temporary drop in morbidity, and how long it would take for another major outbreak to occur.

## Data Availability

All data produced in the present study are available upon reasonable request to the authors.

## Acknowledgements

This study was supported by grants from the Key Program of the National Natural Science Foundation of China (grant 82130093 to H.Y.).

