## supplementary for "Long-term neutralizing antibody dynamics against SARS-CoV-2 in symptomatic and asymptomatic infections: a systematic review and meta-analysis"

#### Supplementary Tables

##### Table S1. Search strategy for three peer-reviewed databases and three preprint servers

| Database | Step | Search strategy | Number of articles* |
|---|---|---|---|
| PubMed | #1 | Title/Abstract: 2019-nCoV OR "coronavirus disease 2019" OR COVID-19 OR "severe acute respiratory syndrome coronavirus 2" OR SARS-CoV-2 | 284,928 |
| | #2 | MeSH Terms: SARS-CoV-2 OR COVID-19 | 223,198 |
| | #3 | #1 OR #2 | 294,777 |
| | #4 | Title/Abstract: "neutralizing antibodies" OR "neutralizing antibody" OR neutraliz* OR "neutralization titer" OR "neutralization titers" OR neutralization | 107,200 |
| | #5 | MeSH Terms: "Antibodies, Neutralizing" | 16,764 |
| | #6 | #4 OR #5 | 110,360 |
| | #7 | Title/Abstract: dynamic* OR kinetic* OR dura* OR persist* OR longitudinal OR follow-up OR waning OR decay OR long-term OR short-term | 4,521,061 |
| | #8 | Title/Abstract: patient* OR outpatient* OR inpatient* OR hospitalized OR non-hospitalized OR convalescent OR convalescence OR recovered OR case* OR infection* OR symptomatic | 10,902,735 |
| | #9 | Title/Abstract: asymptomatic OR inapparent OR subclinical OR presymptomatic | 235,268 |
| | #10 | MeSH Terms: asymptomatic infection | 2,614 |
| | #11 | #9 OR #10 | 235,714 |
| | #12 | #8 OR #11 | 10,945,386 |
| | #13 | Language: English | 29,906,354 |
| | #14 | 2020/01/01 to 2022/10/02 | 4,317,225 |
| | #15 | #3 AND #6 AND #7 AND #12 AND #13 AND #14 | 1,432 |
| Embase | #1 | Title/Abstract: 2019-nCoV OR "coronavirus disease 2019" OR COVID-19 OR "severe acute respiratory syndrome coronavirus 2" OR SARS-CoV-2 | 303,357 |
| | #2 | Title/Abstract: "neutralizing antibodies" OR "neutralizing antibody" OR neutraliz* OR "neutralization titer" OR "neutralization titers" OR neutralization | 128,772 |
| | #3 | Title/Abstract: dynamic* OR kinetic* OR dura* OR persist* OR longitudinal OR follow-up OR waning OR decay OR long-term OR short-term | 6,005,279 |
| | #4 | Title/Abstract: patient* OR outpatient* OR inpatient* OR hospitalized OR non-hospitalized OR convalescent OR convalescence OR recovered OR case* OR infection* OR | 15,027,692 |

| | | | |
|---|---|---|---|
| | | symptomatic | |
| | #5 | Title/Abstract: asymptomatic OR inapparent OR subclinical OR presymptomatic | 342,084 |
| | #6 | #4 OR #5 | 15,081,386 |
| | #7 | Language: English | 35,608,711 |
| | #8 | 2020/01/01 to 2022/10/02 | 5,868,242 |
| | #9 | #1 AND #2 AND #3 AND #6 AND #7 AND #8 | 1,448 |
| Web of Science | #1 | Title/Abstract: 2019-nCoV OR "coronavirus disease 2019" OR COVID-19 OR "severe acute respiratory syndrome coronavirus 2" OR SARS-CoV-2 | 412,852 |
| | #2 | Title/Abstract: "neutralizing antibodies" OR "neutralizing antibody" OR neutraliz* OR "neutralization titer" OR "neutralization titers" OR neutralization | 289,448 |
| | #3 | Title/Abstract: dynamic* OR kinetic* OR dura* OR persist* OR longitudinal OR follow-up OR waning OR decay OR long-term OR short-term | 13,554,316 |
| | #4 | Title/Abstract: patient* OR outpatient* OR inpatient* OR hospitalized OR non-hospitalized OR convalescent OR convalescence OR recovered OR case* OR infection* OR symptomatic | 16,249,972 |
| | #5 | Title/Abstract: asymptomatic OR inapparent OR subclinical OR presymptomatic | 292,190 |
| | #6 | #4 OR #5 | 16,358,081 |
| | #7 | Language: English | 141,064,630 |
| | #8 | 2020/01/01 to 2022/10/02 | 23,521,248 |
| | #9 | #1 AND #2 AND #3 AND #6 AND #7 AND #8 | 1,433 |
| Europe PMC | #1 | Title/Abstract: 2019-nCoV OR "coronavirus disease 2019" OR COVID-19 OR "severe acute respiratory syndrome coronavirus 2" OR SARS-CoV-2 | 383,770 |
| | #2 | Title/Abstract: "neutralizing antibodies" OR "neutralizing antibody" OR neutraliz* OR "neutralization titer" OR "neutralization titers" OR neutralization | 196,307 |
| | #3 | Title/Abstract: dynamic* OR kinetic* OR dura* OR persist* OR longitudinal OR follow-up OR waning OR decay OR long-term OR short-term | 9,859,724 |
| | #4 | Title/Abstract: patient* OR outpatient* OR inpatient* OR hospitalized OR non-hospitalized OR convalescent OR convalescence OR recovered OR case* OR infection* OR symptomatic | 11,459,923 |
| | #5 | Title/Abstract: asymptomatic OR inapparent OR subclinical OR presymptomatic | 238,935 |
| | #6 | #4 OR #5 | 11,502,928 |
| | #7 | Type: Preprints | 494,281 |

| | | | |
|---|---|---|---|
| | #8 | 2020/01/01 to 2022/10/02 | 77,970 |
| | #9 | #1 AND #2 AND #3 AND #6 AND #7 AND #8 | 3,030 |

**Table S2. Inclusion and exclusion criteria**

| | |
|---|---|
| Inclusion criteria | 1) Studies that reported neutralizing antibodies against SARS-CoV-2 by using serum or plasma collected from symptomatic and asymptomatic individuals without vaccination with virologically or serologically-confirmed SARS-CoV-2 infections.<br>2) Studies that reported the neutralizing antibodies titers expressed as reciprocal dilution of serum that neutralizes or inhibits 50% of the virus.<br>3) Studies that reported individual antibody titers in tables or images that could be digitized the data using a digital extraction tool. |
| Exclusion criteria | 1) Studies that were not in humans.<br>2) Studies that investigated immunogenicity among certain population, including pregnant and lactating women, cancer patients, HIV-positive people or other immunosuppressive patients.<br>3) Studies about comparison of different immunoassays.<br>4) Studies without data of interval between onset/diagnosis and sampling.<br>5) Studies with data not available or with inconclusive data and no responses from the authors..<br>6) Abstracts of congress meetings or conference proceedings, study protocols, media news, commentaries, and reviews. |

**Table S3. Scoring system used for evaluation of published reports describing neutralizing antibodies against SARS-CoV- 2**

| Category | Item | Maximum score | Individual score | |
|---|---|---|---|---|
| | | | 0 | 1 |
| Study design | Clear criteria and definition | 1 | No | Clear inclusion and exclusion criteria for study subjects and definition for asymptomatic or asymptomatic infection |
| | Continuous samples of the same individual | 1 | No | Continuous samples of the same individual |
| | Specific strain of infection | 1 | No | Type of infected strain confirmed by article report or gene sequencing |
| | Comprehensive information about the individuals | 1 | No | Comprehensively collection of the information about the individuals (age, sex, clinical severity, comorbidities, etc.) |
| | Sample size and follow time | 1 | No | Study participants ≥30 and follow-up time ≥30 days |
| Laboratory method | Validated neutralization assay | 1 | No | Use a validated neutralization assay protocol |
| | Details of neutralization assays | 1 | No | Detailed description of sample, virus, and cell types in neutralization assays |
| Outcome | Clear definition of sampling time | 1 | No | Clear definition of sampling time (time since confirmation or symptom onset) |
| | Raw data on individual antibody titers | 1 | No | Providing raw data on individual antibody titers |
| | Total | 9 | | |

### A score of 0–3 was considered as a Low quality study (C), a score of 4–6 was considered as a Moderate quality study (B), and a score of 7–9 was considered as a High quality study (A).

### Table S4. Summary of characteristics of included studies

| Author (Year, Location) | Study participants | | | Laboratory methods | | | | | | | Outcome | | |
|---|---|---|---|---|---|---|---|---|---|---|---|---|---|
| | Participants (N) | Age (Median, range/mean±SD) | Sampling time | Serological assay | Vector type | Type of Samples | Dilution factor (Dilution range) | Fold | Virus | Cell line | Outcome variable | Geometric mean titer (95%CI) | Note |
| Prototype strain | | | | | | | | | | | | | |
| Lau et al. (2021, China) | Asymptomatic (31) | Range: ≥15 | 76 days after confirmation | PRNT | - | Serum | 1:10 – 1:320 | - | BetaCoV/HongKong/VM20001061/2020 | Vero E6 cells | PRNT50 | - | - |
| | Mild (151) | All ages | 209 days after symptom onset | | | | | | | | | | |
| | Severe (13) | Range: ≥16 | | | | | | | | | | | |
| Noh et al. (2021, Korea) | Asymptomatic (14) | Mean (SD): 51.3 (10.4) | 8 weeks after diagnosis | PRNT | - | Serum | 1:20 - 1:20480 | 4-fold | BetaCoV/Korea/KCDC03/2020 | Vero E6 cells | PRNT50 | 373.24 | Extract individual data from plot |
| | | Mean (SD): 44.7 (8.9) | 20 weeks after diagnosis | | | | | | | | | 114.58 (53.65–244.70) | |
| | | Mean (SD): 47.3 (8.0) | 27 weeks after diagnosis | | | | | | | | | 100.34 (15.35–655.77) | |
| | Mild (42) | Mean (SD): 44.7 (8.9) | 8 weeks after diagnosis | | | | | | | | | 232.237 | |
| | | Mean (SD): 44.7 (8.9) | 20 weeks after diagnosis | | | | | | | | | 166.47 (127.27–217.73) | |

| | | | | | | | | | | | | | |
|---|---|---|---|---|---|---|---|---|---|---|---|---|---|
| | | Mean (SD): 44.7 (8.9) | 27 weeks after diagnosis | | | | | | | | | 64.85 (40.17–104.70) | |
| | Severe (41) | Mean (SD): 47.3 (8.0) | 8 weeks after diagnosis | | | | | | | | | 1054.33 | |
| | | Mean (SD): 47.3 (8.0) | 20 weeks after diagnosis | | | | | | | | | 249.15 (181.54–341.95) | |
| | | Mean (SD): 47.3 (8.0) | 27 weeks after diagnosis | | | | | | | | | 158.55 (98.82–254.40) | |
| Gaebler et al. (2021, USA) | Mild (76), Severe (10) | - | 6.2 months after symptom onset | Pseudotyped virus neutralization assay | Lentivirus | Plasma | - | 4-fold | SARS-CoV-2 pseudotyped virus | 293TACE2 cells | NT50 | - | - |
| Garcia-Beltran et al. (2021, USA) | Symptomatic (98) | - | 60 days after symptom onset | Pseudotyped virus neutralization assay | Lentivirus | Serum | 1:12 - 1:8748 | 3-fold | - | 293T-ACE2 cells | NT50 | - | Extract individual data from plot |
| Yamayoshi et al. (2021, Japan) | Mild (13), Severe (26) | - | 154 days after symptom onset | PRNT | - | Serum or plasma | 1:10- | 3-fold | - | Vero-TMPRSS2 cells | PRNT50 | - | Extract individual data from plot |
| Wheatley et al. (2021, Australia) | Asymptomatic (1) | - | 113 days after confirmation | Microneutralisation assays | - | Plasma | 1:20 - 1:10240 | - | SARS-CoV-2 isolate CoV/Australia/VIC01/2020 | Vero cells | NT50 | - | - |
| | Symptomatic (63) | | 149 days after symptom onset | | | | | | | | | | |
| Padoan et al. (2021, Italy) | Severe (62) | - | 93 days after symptom onset | PRNT | - | Serum | - | 2-fold | - | Vero E6 cells | PRNT50 | - | - |
| Yang et al. (2022, China) | Asymptomatic (29) | Median (range): 25 (7-58) | 480 days after confirmation | Microneutralisation assays | - | Plasma | 1:10 - 1:5120 | 2-fold | BetaCoV/Shenzhen/SZTH-003/2020 | Vero E6 cells | NT50 | - | - |

| | | | | | | | | | | | | | |
|---|---|---|---|---|---|---|---|---|---|---|---|---|---|
| | Mild (134) | Median (range): 43 (2-79) | | | | | | | (EPI_ISL_406594) | | | | |
| | Severe (51) | Median (range): 62 (31-76) | | | | | | | | | | | |
| Wang et al.1 (2020, China) | Mild (11), Severe (12) | Median (range): 56 (24–82) | 42 days after illness onset | FRNT | - | Plasma | - | - | Authentic SARS-CoV-2 virus | Vero E6 cells | FRNT50 | - | Extract individual data from plot |
| Glans et al. (2021, Sweden) | Severe (36) | Median (range): 60.5 (25–77) | 53 days after symptom onset | Microneutralization assays | - | Serum | 1:10 - 1:10240 | 2-fold | - | Vero E6 cells | NT50 | - | - |
| Vicenti et al. (2021, Italy) | Asymptomatic (42) | Median (IQR): 50 (39–57) | 89 days after diagnosis | Live virus neutralization assay | - | Serum | 1:4 - 1:512 | 2-fold | SARS-CoV-2 lineage B | Vero E6 cells | EC50 | Median (IQR): 15.5 (8.5–37) | Extract individual data from plot |
| | | | 268 days after diagnosis | | | | | | | | | Median (IQR): 13 (2–29) | |
| | Mild (25) | Median (IQR): 49 (46–55) | 84 days after diagnosis | | | | | | | | | Median (IQR): 57.5 (15.1–121) | |
| | | | 225 days after diagnosis | | | | | | | | | Median (IQR): 31 (13.6–57.1) | |
| Suthar et al. (2020, USA) | Symptomatic (44) | Range: 25-87 | 30 days after symptom onset | FRNT | - | Serum or plasma | - | - | SARS-CoV-2 (2019-nCoV/USA_WA1/2020) | Vero E6 cells | FRNT50 | - | - |

| | | | | | | | | | | | | | |
|---|---|---|---|---|---|---|---|---|---|---|---|---|---|
| Wang et al.2 (2020, China) | Symptomatic (8) | - | 60 days after symptom onset | Modified cytopathogenic assay | - | Serum | - | 2-fold | - | Vero cells | NT50 | - | Extract individual data from plot |
| Bal et al. (2021, France) | Mild (57) | Median: 70.4 | 14 days after symptom onset | PRNT | - | Serum | - | 2-fold | wild type SARS-CoV-2 | Vero E6 cells | PRNT50 | Median (IQR): 60 (40-100) | Extract individual data from plot |
| | | | 28 days after symptom onset | | | | | | | | | Median (IQR): 80 (60-120] | |
| | | | 89 days after symptom onset | | | | | | | | | Median (IQR): 60 (40-120) | |
| | Severe (44) | | 14 days after symptom onset | | | | | | | | | Median (IQR): 160 (80-320) | |
| | | | 28 days after symptom onset | | | | | | | | | Median (IQR): 480 (240-640) | |
| | | | 89 days after symptom onset | | | | | | | | | Median (IQR): 320 (120-640) | |
| Cohen et al. (2021, USA) | Symptomatic (183) | - | 250 days after symptom onset | FRNT | - | Serum or plasma | - | 3-fold | 2019-nCoV/USA_WA1/2020 strain | Vero E6 cells | FRNT50 | - | Extract individual data from plot |
| Flehmig et al. (2020, Germany) | Mild (3) | - | 193 days after symptom onset | Virus neutralization assay | - | Serum | 1:40 - 1:5120 | 2-fold | icSARS-CoV-2-mNG | human cells Caco-2 (human colorectal adenocarcinoma) | ID50 | - | Extract individual data from plot |

| | | | | | | | | | | | | | |
|---|---|---|---|---|---|---|---|---|---|---|---|---|---|
| Deshpande et al. (2020, India) | Symptomatic (12) | - | 25 days after symptom onset | Microneutralisation assay | - | Serum | - | 2-fold | Vero CCL-81-adapted SARS-CoV-2 Indian isolate virus | Vero CCL-81 cells | NT50 | - | Extract individual data from plot |
| Ramos et al. (2021, USA) | Asymptomatic (11)<br>Mild (27) | - | 84 days after outbreak | Live virus neutralization assay | - | Serum | 1:10 - | 2-fold | MNeonGreen SARS-CoV-2 | Vero E6 cells | ID50 | - | - |
| Sun et al. (2020, China) | Mild (28), Severe (7) | - | 40 days after illness onset | Microneutralisation assay | - | Serum | 1:4 - 1:1024 | 2-fold | SARS-CoV-2 (no. EPI_ISL_403934) | Vero E6 cells | NT50 | - | Extract individual data from plot |
| Dufloo et al. (2021, France) | Mild (21) | - | 77 days after symptom onset | Real-virus neutralization | - | Serum | - | - | BetaCoV/France/IDF0372/2020 | U2OS-ACE2-GFP-1-10+U2OS-ACE2-GFP-11 cells | ID50 | - | - |
| Trinité et al. (2021, Spain) | Symptomatic (40) | Median (range): 63 (56–70) | 73 days after symptom onset | Live virus neutralization assay | - | Plasma | - | - | SARS-CoV-2 isolate Cat01 (accession ID EPI_ISL_418268) | Vero E6 cells | IC50 | - | Extract individual data from plot |
| Vanshylla et al. (2021, Germany) | Asymptomatic (44) | - | Median 7.3 weeks after infection | Pseudovirus neutralizing assay | Lentivirus vector | Serum | - | - | SARS-CoV-2 Wu01 spike (EPI_ISL_406716) | 293T cells | ID50 | - | Extract individual data from plot |
| Wang et al.3 (2020, China) | Symptomatic (30) | Median (IQR): 52 (45-67) | 3 months after symptom onset | Pseudovirus neutralizing assay | VSV vector | Serum | - | - | SARS-CoV-2 S protein (QHD43416) | 293T-ACE2 cells | ID50 | - | Extract individual data from plot |

| | | | | | | | | | | | | | |
|---|---|---|---|---|---|---|---|---|---|---|---|---|---|
| Lei et al. (2020, China) | Asymptomatic (36) | Median (IQR): 46(30-59) | 65 days after exposure | Pseudovirus neutralizing assay | Lentivirus vector | Serum | 1:10-1:2560 | 2-fold | - | Vero E6 cells | NT50 | Median (IQR): 14.17 (1.97-25.22) | Extract individual data from plot |
| | Mild (51) | | | | | | | | | | | Median (IQR): 242.32 (38.50-796.20) | |
| Moriyama et al. (2021, Japan) | Mild (59), Severe (129) | - | 10 months after symptom onset | Pseudovirus neutralization assay | VSV vector | Plasma | 1:10- | 5-fold | SARS-CoV-2 pseudovirus | VeroE6/TMPRSS2 cells | IC50 | - | Extract individual data from plot |
| Pradenas et al. (2021, Spain) | Severe (104) | Median (IQR): 57.5 (46, 66) | 242 days after symptom onset | Pseudovirus neutralization assay | Lentivirus vector | Plasma | 1:60 - 1:14580 | 3 | HIV reporter pseudoviruses expressing SARS-CoV-2 S protein and Luciferase | HEK293T cells overexpressing WT human ACE-2 | ID50 | - | Extract individual data from plot |
| Sakharkar et al. (2021, USA) | Mild (5), Severe (3) | - | 168 days after illness or positive test | Microneutralization assays | VSV vector | Serum | - | 2 | VSV based pseudovirus | 293T-hsACE2 cells | NT50 | - | - |
| Tan et al. (2020, China) | Symptomatic (22) | All ages | 7 months after symptom onset | Pseudovirus-based SARS-CoV-2 neutralizing assay | VSV vector | Plasma | - | - | SARS-CoV-2 | Huh7 cells | NT50 | - | - |
| Crawford et al. (2020, USA) | Asymptomatic (6) | Median (range): 64 (24–79) | 131 days after confirmation | Pseudovirus neutralizing assay | Lentivirus vector | Plasma | 1:20-1:14580 | 3-fold | Wuhan/Hu-1/2019 | 293T-ACE2 cells | NT50 | - | - |
| | Mild (21) | Median (range): 43 (22–76) | 152 days after symptom onset | | | | | | | | | | |

| | | | | | | | | | | | | | |
|---|---|---|---|---|---|---|---|---|---|---|---|---|---|
| | Severe (5) | Median (range): 54 (31–64) | 121 days after symptom onset | | | | | | | | | | |
| Dispinseri et al.1(2021, Italy) | Symptomatic (162) | Median(95%CI): 63 (52–72.5) | 2 weeks after symptom onset | Pseudovirus neutralization assay | Lentivirus vector | Serum | 1:40- | 3-fold | - | Vero E6 cells | ID50 | Mean (SD): 3500 (8600) | Extract individual data from plot |
| | | | 4 weeks after symptom onset | | | | | | | | | Mean (SD): 45300 (252000) | |
| | | | 8 weeks after symptom onset | | | | | | | | | Mean (SD): 9950 (24400) | |
| | | | 16 weeks after symptom onset | | | | | | | | | Mean (SD): 2160 (3910) | |
| | | | 36 weeks after symptom onset | | | | | | | | | Mean (SD): 1460 (1910) | |
| Dispinseri et al.2 (2021, Italy) | Severe (110) | - | 38 days after symptom onset | Pseudovirus neutralization assay | Lentivirus vector | Serum | - | - | - | Vero E6 cells | ID50 | Median (IQR): 3593 (1753-6419) | Extract individual data from plot |
| Harrington et al. (2021, USA) | Symptomatic (33) | Median (range): 41 (24–74) | 6 months after symptom onset | Pseudovirus neutralization assay | Lentivirus vector | Plasma | 1:50-1:109350 | - | - | 293T/hACE2 cells | ID50 | - | Extract individual data from plot |
| Hashem et al. (2020, Saudi Arabia) | Mild (46), Severe (41) | - | 70 days after symptom onset | Pseudovirus neutralization assay | VSV vector | Serum | 1:20- | 2-fold | - | Vero E6 cells | IC50 | - | Extract individual data from plot |
| Dan et al. (2021, USA) | Mild (170), Severe (13) | - | 240 days after symptom onset | Pseudovirus neutralization assay | VSV vector | Plasma | 1:20- | - | Wuhan-Hu-1 | Vero E6 cells | IC50 | - | - |
| Iyer et al. (2020, USA) | Symptomatic (15) | Median (IQR): 59 (45–71) | 122 days after symptom onset | Pseudovirus neutralization assay | Lentivirus vector | Plasma | - | 2-fold | - | HEK293T-hACE2 cells | NT50 | - | - |

| | | | | | | | | | | | | | |
|---|---|---|---|---|---|---|---|---|---|---|---|---|---|
| Anand et al. (2021, Canada) | Mild (32) | Median (range): 47 (20-65) | 95 days after symptom onset | Pseudovirus neutralization assay | Lentivirus vector | Plasma | 1:50-1:31250 | 5-fold | - | 293T-ACE2 cells | ID50 | 155.33 | Extract individual data from plot |
| | Mild (28) | Median (range): 47 (20-65) | 127 days after symptom onset | | | | | | | | | 57.76 | |
| | Mild (28) | Median (range): 48 (20-65) | 171 days after symptom onset | | | | | | | | | 53.956 | |
| | Mild (13) | Median (range): 46 (20-65) | 233 days after symptom onset | | | | | | | | | 61.833 | |
| Yao et al. (2021, China) | Symptomatic (34) | Median (IQR): 52 (38-67) | 98 days after illness onset | Pseudovirus-based neutralization assay | VSV vector | Serum | 1:10- | 2-fold | VSV-SARS-CoV-2-Sdel18 virus | BHK21-hACE2 cells | NT50 | - | Extract individual data from plot |
| Bastug et al. (2022, Turkey) | Mild (23) | Mean (SD): 37.7 (10.4) | 10 months after symptom onset | Live virus neutralization assay | - | Serum | 1:5- | 2-fold | SARS-CoV-2 Ank1 isolate | Vero E6 cells | NT50 | - | Extract individual data from plot |
| | Symptomatic (19) | Mean (SD): 42.7 (9.6) | | | | | | | | | | | |
| Dupont et al. (2021, UK) | Severe (20) | Median (IQR): 50 (23-83) | 305 days after symptom onset | Pseudovirus neutralization assay | Lentivirus vector | Serum | - | - | Wuhan-1 spike protein (WT) | Hela cells | ID50 | - | - |
| Gniadek et al. (2021, USA) | Mild (47) | - | 73 days after symptom onset | Fluorescent SARS-CoV-2 neutralization | - | Plasma | 1:20 - 1:5120 | 2-fold | mNeonGreen SARS-CoV-2 | Vero E6 cells | NT50 | - | Extract individual data from plot |

| | | | | | | | | | | | | | |
|---|---|---|---|---|---|---|---|---|---|---|---|---|---|
| Souza et al. (2021, Brazil) | Asymptomatic (2) | Median (IQR): 34 (31-43) | 102 days after symptom onset | Live virus neutralization assay | - | Plasma | 1:20 - 1:2560 | 2-fold | lineage B (isolate SARS.CoV2/ SP02.2020) | Vero cell | NT50 | - | - |
| | Symptomatic (19) | | 71 days after confirm | | | | | | | | | | |
| Ruetalo et al. (2021, Germany) | Symptomatic (3) | - | 423 days after symptom onset | Live virus neutralization assay | - | Serum | 1:40 - 1:5120 | 2-fold | icSARS-CoV-2-mNG | Caco-2 cells | NT50 | - | Extract individual data from plot |
| Lei et al. (2021, China) | Symptomatic (42) | - | 60 days after symptom onset | Pseudotyped virus neutralization assay | Lentivirus vector | Serum | 1:10-1:2560 | 2-fold | SARS-CoV-2 pseudotyped virus | Vero E6 cells | NT50 | - | Extract individual data from plot |
| Vacharathit et al. (2021, Thailand) | Mild (23) | Median (IQR):34.8 (27.0-39.9) | 12 months after symptom onset | Microneutralization assays | - | Serum | 1:10- | 2-fold | SARS-CoV-2/01/human/Jan2020/Thailand | Vero E6 cells | NT50 | - | Extract individual data from plot |
| | Severe (52) | - | | | | | | | | | | | |
| Selhorst et al. (2021, Belgium) | Asymptomatic (1) | 87 | 12 days after diagnosis | Live virus neutralization assay | - | Serum | 1:50-1:1600 | - | 2019-nCoV-Italy-INMI1 | Vero E6 cells | NT50 | - | - |
| | Mild (3) | Range: 39-77 | 94 days after symptom onset | | | | | | | | | | |
| Choe et al. (2022, Korea) | Asymptomatic (4) | Range: 20-26 | 12 months after symptom onset/confirmation | Microneutralization assays | - | Serum | 1:20-1:12500 | 5-fold | SARS-CoV-2/wt (NCCP No. 43326) | - | FRNT50 | - | Extract individual data from plot |
| | Severe (12) | Range: 39-76 | | | | | | | | | | | |
| Pradenas et al. (2022, Spain) | Severe (84) | Median (IQR):58 (48-67) | Median (IQR): 185 (5–266) days after symptom onset | Pseudovirus neutralization assay | Lentivirus vector | Plasma | 1:60-1:14580 | 3-fold | SARS-CoV-2.SctD19 (WH1) | HEK293T/ hACE2 cells | NT50 | - | Extract individual data from plot |
| Lee et al. (2021, Singapore) | Mild (42) | - | Median 98 days after illness onset | Pseudovirus neutralization assay | Lentivirus vector | Plasma | 1:10 - 1:31250 | - | SARS-CoV-2 with wildtype S | CHO-ACE2 cells | EC50 | - | - |
| | Severe (8) | | | | | | | | | | | | |

| | | | | | | | | | | | | | |
|---|---|---|---|---|---|---|---|---|---|---|---|---|---|
| Ma et al. (2022, China) | Asymptomatic (30) | Median (IQR): 30.5 (26.3-33.0) | 5 days after the first positive RT-PCR results | Pseudovirus neutralization assay | Lentivirus vector | Serum | - | - | - | Huh7/ACE 14 cells | ID50 | Median (IQR): 586.1 (397.6-886.7) | Extract individual data from plot |
| | | | 10 days after the first positive RT-PCR results | | | | | | | | | Median (IQR): 346.5 (206.6-566.5) | |
| | | | 29 days after the first positive RT-PCR results | | | | | | | | | Median (IQR): 62.0 (20.9-183.8） | |
| | | | 53 days after the first positive RT-PCR results | | | | | | | | | Median (IQR): 125.2 (70.9-480.8) | |
| | | | 107 days after the first positive RT-PCR results | | | | | | | | | Median (IQR): 345.5 (143.4-811.4) | |
| | Symptomatic (20) | Median (IQR): 30.5 (29.0-32.8) | 5 days after symptom onset | | | | | | | | | Median (IQR): 29.2 (15.9- 6.9) | |
| | | | 10 days after symptom onset | | | | | | | | | Median (IQR): 189.4 (87.1-411.9) | |
| | | | 16 days after symptom onset | | | | | | | | | Median (IQR): 1011.5 (564.9-1862.3) | |
| | | | 29 days after symptom onset | | | | | | | | | Median (IQR): 1855.1 | |

| | | | | | | | | | | | | | |
|---|---|---|---|---|---|---|---|---|---|---|---|---|---|
| | | | | | | | | | | | | (1190.1-2812.9) | |
| | | | 107 days after symptom onset | | | | | | | | | Median (IQR): 858.0 (492.6-1453.5) | |
| Alpha strain | | | | | | | | | | | | | |
| Dupont et al. (2021, UK) | Severe (32) | Median (IQR): 63 (37–96) | 80 days after symptom onset | Pseudovirus neutralization assay | Lentivirus vector | Serum | - | - | B.1.1.7 spike | HeLa cells stably expressing the ACE2 receptor | ID50 | - | - |
| Pradenas et al. (2022, Spain) | Severe (22) | Median (IQR):56 (49–62) | Median (IQR): 26 (16–76) days after symptom onset | Pseudovirus neutralization assay | Lentivirus vector | Plasma | 1:60-1:14580 | 3-fold | SARS-CoV-2.SctD19 (alpha) | HEK293T/ hACE2 cells | NT50 | - | Extract individual data from plot |
| Omicron BA.2 strain | | | | | | | | | | | | | |
| Cheng et al. (2022, China) | Asymptomatic (2) | Range: 61-70 | 44 days after first RT-PCR positive test | PRNT | - | Serum | 1:10-1:320 | 2-fold | BA.2(hCoV-19/Hong Kong/VM22000135_HKUVOC0588P2/2022) | Vero E6 TMPRSS2 cells | PRNT50 | - | - |
| | Mild (5) | Range: ≥51 | 52 days after symptom onset | | | | | | | | | | |

Abbreviations: PRNT, Plaque reduction neutralization test; FRNT, Focus reduction neutralization assays.

#### Table S5. Quality assessment of studies

| Reference | Study design | | | | | Laboratory method | | Outcome | | Total Score | Grade |
|---|---|---|---|---|---|---|---|---|---|---|---|
| | Clear criteria and definition | Continuous samples of the same individual | Specific strain of infection | Comprehensive information about the individuals | Sample size and follow time | Validated neutralization assay | Details of neutralization assays | Clear definition of sampling time | Raw data on individual antibody titers | | |
| Lau et al., 2021 | 1 | 1 | 1 | 1 | 1 | 1 | 1 | 0 | 1 | 8 | A |
| Noh et al., 2021 | 1 | 1 | 1 | 0 | 1 | 1 | 1 | 0 | 0 | 6 | B |
| Gaebler et al., 2021 | 1 | 1 | 1 | 1 | 1 | 0 | 1 | 1 | 1 | 8 | A |
| Garcia-Beltran et al., 2021 | 1 | 0 | 1 | 0 | 1 | 1 | 1 | 1 | 0 | 6 | B |
| Yamayoshi et al., 2021 | 1 | 1 | 1 | 1 | 1 | 0 | 1 | 0 | 0 | 6 | B |
| Wheatley et al., 2021 | 1 | 1 | 1 | 0 | 1 | 0 | 1 | 1 | 1 | 7 | A |
| Padoan et al., 2020 | 1 | 0 | 1 | 1 | 1 | 0 | 1 | 1 | 1 | 7 | A |
| Yang et al., 2022 | 1 | 1 | 1 | 1 | 1 | 1 | 1 | 0 | 1 | 8 | A |
| Wang et al., 2020 | 1 | 1 | 1 | 1 | 1 | 0 | 1 | 0 | 0 | 6 | B |
| Glans et al., 2021 | 1 | 0 | 1 | 0 | 1 | 1 | 1 | 1 | 1 | 7 | A |
| Vicenti et al., 2021 | 1 | 1 | 1 | 0 | 1 | 0 | 1 | 0 | 0 | 5 | B |
| Suthar et al., 2020 | 0 | 0 | 1 | 1 | 1 | 0 | 1 | 1 | 1 | 6 | B |

| | | | | | | | | | | | |
|---|---|---|---|---|---|---|---|---|---|---|---|
| Wang et al., 2020 | 1 | 1 | 1 | 0 | 0 | 0 | 1 | 1 | 0 | 5 | B |
| Bal et al., 2021 | 1 | 1 | 1 | 0 | 1 | 1 | 1 | 1 | 0 | 7 | A |
| Cohen et al., 2021 | 1 | 1 | 1 | 0 | 1 | 1 | 1 | 1 | 0 | 7 | A |
| Flehmig et al., 2020 | 1 | 1 | 1 | 1 | 0 | 0 | 1 | 1 | 0 | 6 | B |
| Deshpande et al., 2020 | 0 | 1 | 1 | 0 | 0 | 1 | 1 | 0 | 0 | 4 | B |
| Ramos et al., 2021 | 1 | 1 | 1 | 0 | 0 | 0 | 1 | 0 | 1 | 5 | B |
| Sun et al., 2020 | 1 | 1 | 1 | 0 | 1 | 0 | 1 | 0 | 0 | 5 | B |
| Dufloo et al., 2021 | 0 | 1 | 1 | 0 | 1 | 0 | 1 | 1 | 1 | 6 | B |
| Trinité et al., 2021 | 1 | 1 | 1 | 0 | 1 | 0 | 1 | 1 | 0 | 6 | B |
| Vanshylla et al., 2021 | 0 | 1 | 1 | 0 | 1 | 0 | 1 | 0 | 0 | 4 | B |
| Wang et al., 2020 | 1 | 1 | 1 | 1 | 1 | 0 | 1 | 1 | 0 | 7 | A |
| Lei et al., 2020 | 1 | 0 | 1 | 0 | 1 | 0 | 1 | 0 | 0 | 4 | B |
| Moriyama et al., 2021 | 1 | 0 | 1 | 0 | 1 | 0 | 1 | 1 | 0 | 5 | B |
| Pradenas et al., 2021 | 1 | 1 | 1 | 0 | 1 | 1 | 1 | 1 | 0 | 7 | A |
| Sakharkar et al., 2021 | 0 | 1 | 1 | 1 | 0 | 1 | 1 | 0 | 1 | 6 | B |
| Tan et al., 2020 | 0 | 0 | 1 | 1 | 0 | 1 | 1 | 1 | 1 | 6 | B |

| | | | | | | | | | | | |
|---|---|---|---|---|---|---|---|---|---|---|---|
| Crawford et al., 2020 | 0 | 1 | 1 | 1 | 1 | 1 | 1 | 1 | 1 | 8 | A |
| Dispinseri et al., 2021 | 1 | 0 | 1 | 0 | 1 | 0 | 0 | 1 | 0 | 4 | B |
| Dispinseri et al., 2021 | 1 | 0 | 1 | 0 | 1 | 0 | 0 | 1 | 0 | 4 | B |
| Harrington et al., 2021 | 0 | 0 | 1 | 0 | 1 | 0 | 0 | 1 | 0 | 3 | C |
| Hashem et al., 2020 | 1 | 1 | 1 | 0 | 1 | 1 | 0 | 1 | 0 | 6 | B |
| Dan et al., 2021 | 1 | 1 | 1 | 1 | 1 | 1 | 1 | 1 | 1 | 9 | A |
| Iyer et al., 2020 | 0 | 0 | 1 | 0 | 1 | 1 | 0 | 1 | 1 | 5 | B |
| Anand et al., 2021 | 0 | 1 | 1 | 0 | 1 | 0 | 0 | 1 | 0 | 4 | B |
| Yao et al., 2021 | 0 | 1 | 1 | 0 | 1 | 1 | 1 | 0 | 0 | 5 | B |
| Bastug et al., 2022 | 1 | 1 | 1 | 1 | 1 | 0 | 1 | 1 | 0 | 7 | A |
| Dupont et al., 2021 | 1 | 1 | 1 | 0 | 1 | 0 | 1 | 1 | 1 | 7 | A |
| Gniadek et al., 2021 | 1 | 0 | 1 | 0 | 1 | 0 | 1 | 1 | 0 | 5 | B |
| Souza et al., 2021 | 1 | 0 | 1 | 1 | 0 | 1 | 1 | 1 | 1 | 7 | A |
| Ruetalo et al., 2021 | 0 | 1 | 1 | 1 | 0 | 0 | 1 | 1 | 0 | 5 | B |
| Lei et al., 2021 | 1 | 1 | 1 | 0 | 1 | 1 | 1 | 1 | 0 | 7 | A |
| Vacharathit et al., 2021 | 1 | 1 | 1 | 0 | 1 | 0 | 1 | 0 | 0 | 5 | B |

| | | | | | | | | | | | |
|---|---|---|---|---|---|---|---|---|---|---|---|
| Selhorst et al., 2021 | 1 | 0 | 1 | 1 | 0 | 0 | 1 | 1 | 1 | 6 | B |
| Choe et al., 2022 | 1 | 1 | 1 | 0 | 0 | 0 | 0 | 0 | 0 | 3 | C |
| Pradenas et al., 2022 | 0 | 1 | 1 | 0 | 1 | 1 | 1 | 1 | 0 | 6 | B |
| Cheng et al., 2022 | 0 | 1 | 1 | 1 | 0 | 1 | 1 | 1 | 1 | 7 | A |
| Lee et al., 2021 | 1 | 1 | 1 | 0 | 1 | 1 | 1 | 0 | 1 | 7 | A |
| Ma et al., 2022 | 1 | 1 | 1 | 1 | 1 | 0 | 0 | 1 | 0 | 6 | B |

**Table S6. Univariate analysis and multivariable analysis**

| Variables | No (%) of samples included in analysis | Univariate analysis Coefficients | P | No (%) of samples included in analysis | Multivariable analysis Coefficients | P |
|---|---|---|---|---|---|---|
| / | / | / | / | / | 72.7 | < 0.001*** |
| Sex | 1861 | | | | | |
| (Intercept) | / | 169.2 | < 0.001*** | / | / | / |
| Male | 1112 (59.8%) | Ref | Ref | / | Ref | Ref |
| Female | 749 (40.2%) | 149.3 | 0.102 | / | / | / |
| Age | 1653 | | | 1326 | | |
| (Intercept) | / | 137.6 | < 0.001*** | / | / | / |
| 15-60y | 1129 (68.3%) | Ref | Ref | 898 (67.7%) | Ref | Ref |
| ≤15y | 54 (3.3%) | 47.9 | < 0.001*** | 54 (4.1%) | 52.1 | 0.14 |
| >60y | 470 (30.69%) | 209.2 | < 0.001*** | 374 (28.2%) | 84.4 | 0.14 |
| Comorbidity | 721 | | | / | | |
| (Intercept) | / | 100.7 | < 0.001*** | / | / | / |
| Without comorbidity | 440 (61.0%) | Ref | Ref | / | Ref | Ref |
| With comorbidity | 281 (39.0%) | 91.8 | 0.429 | / | / | / |
| Clinical severity | 3837 | | | 1326 | | |
| (Intercept) | / | 104.2 | < 0.001*** | / | / | / |
| Mild | 1706 (44.5%) | Ref | Ref | 842 (63.5%) | Ref | Ref |
| Severe | 1746 (45.5%) | 509.0 | < 0.001*** | 415 (31.3%) | 200.2 | < 0.001*** |
| Asymptomatic | 385 (10.0%) | 65.8 | < 0.001*** | 69 (5.2%) | 60.1 | 0.34 |
| Assays | 5546 | | | 1326 | | |
| (Intercept) | / | 112.6 | < 0.001*** | / | / | / |
| Live | 2455 (44.3%) | Ref | Ref | 818 (61.7%) | Ref | Ref |
| Lentivirus | 2001 (36.1%) | 469.3 | < 0.001*** | 261 (19.7%) | 148.2 | < 0.001*** |
| VSV | 1090 (19.7%) | 332.2 | < 0.001*** | 247 (18.6%) | 107.4 | 0.0006 *** |

#### Appendix figures

##### Figure S1. Quality scores assigned to studies by clinical categories

(A) Median quality score and range from assessment of studies of symptomatic and asymptomatic individuals. (B) Quality of studies by grade category (i.e., A, B and C). Category A included studies with scores ranging from 7 to 9, category B from 4 to 6 and category C from 0 to 3.

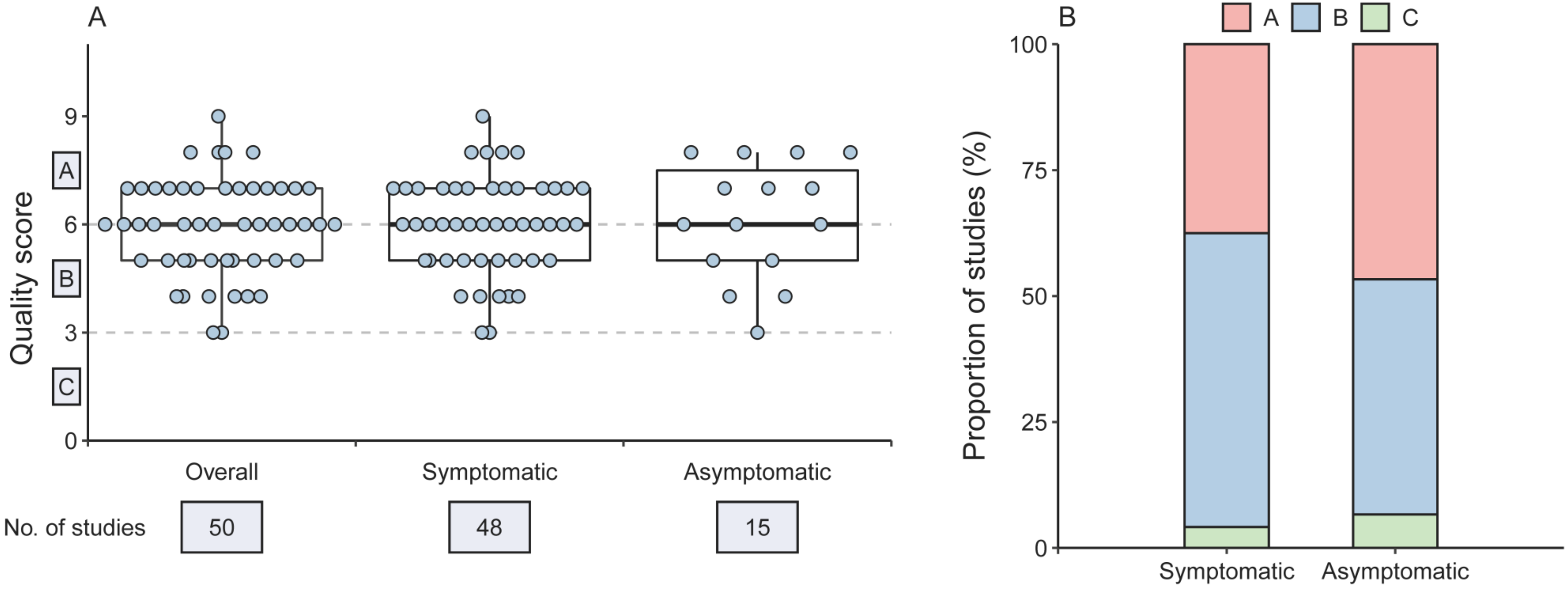

**Figure S2.** **Neutralization antibody responses in prototype strain infections in live virus neutralization assays (all samples)**

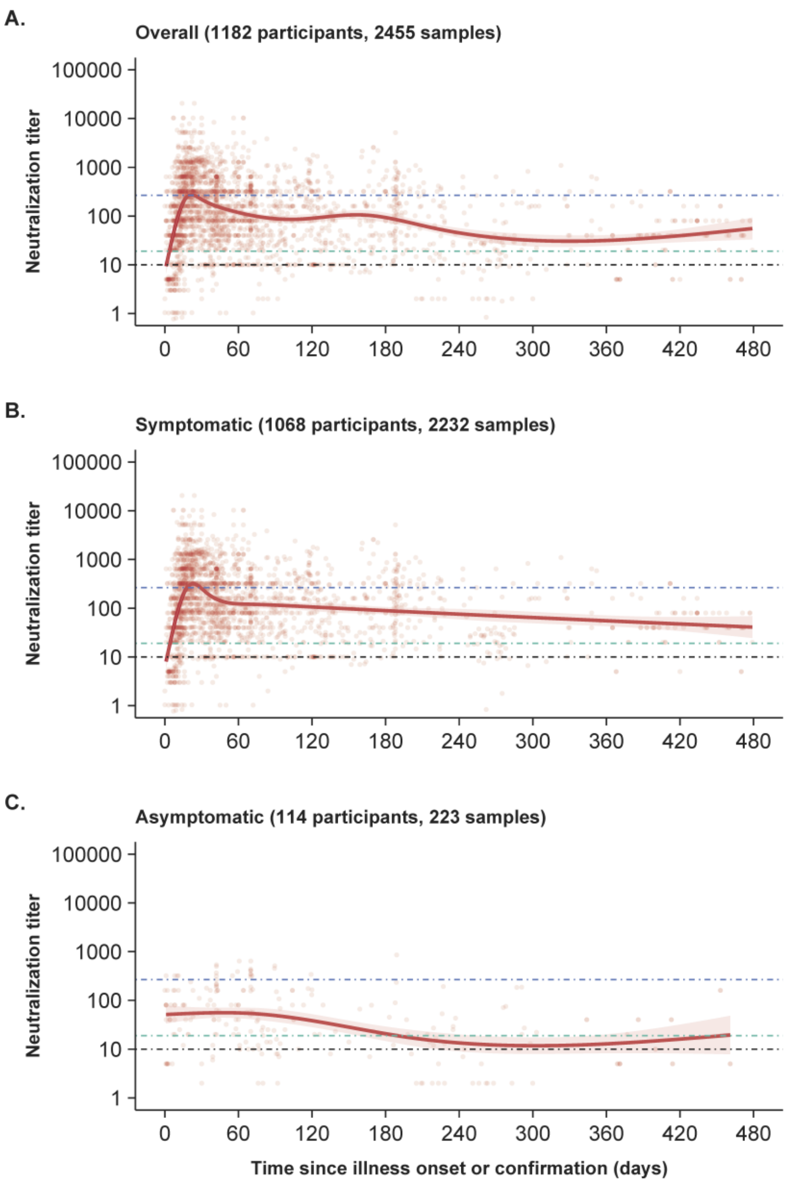

**Figure S3. Comparison of neutralization antibody responses between prototype strain and VOCs (all samples)**

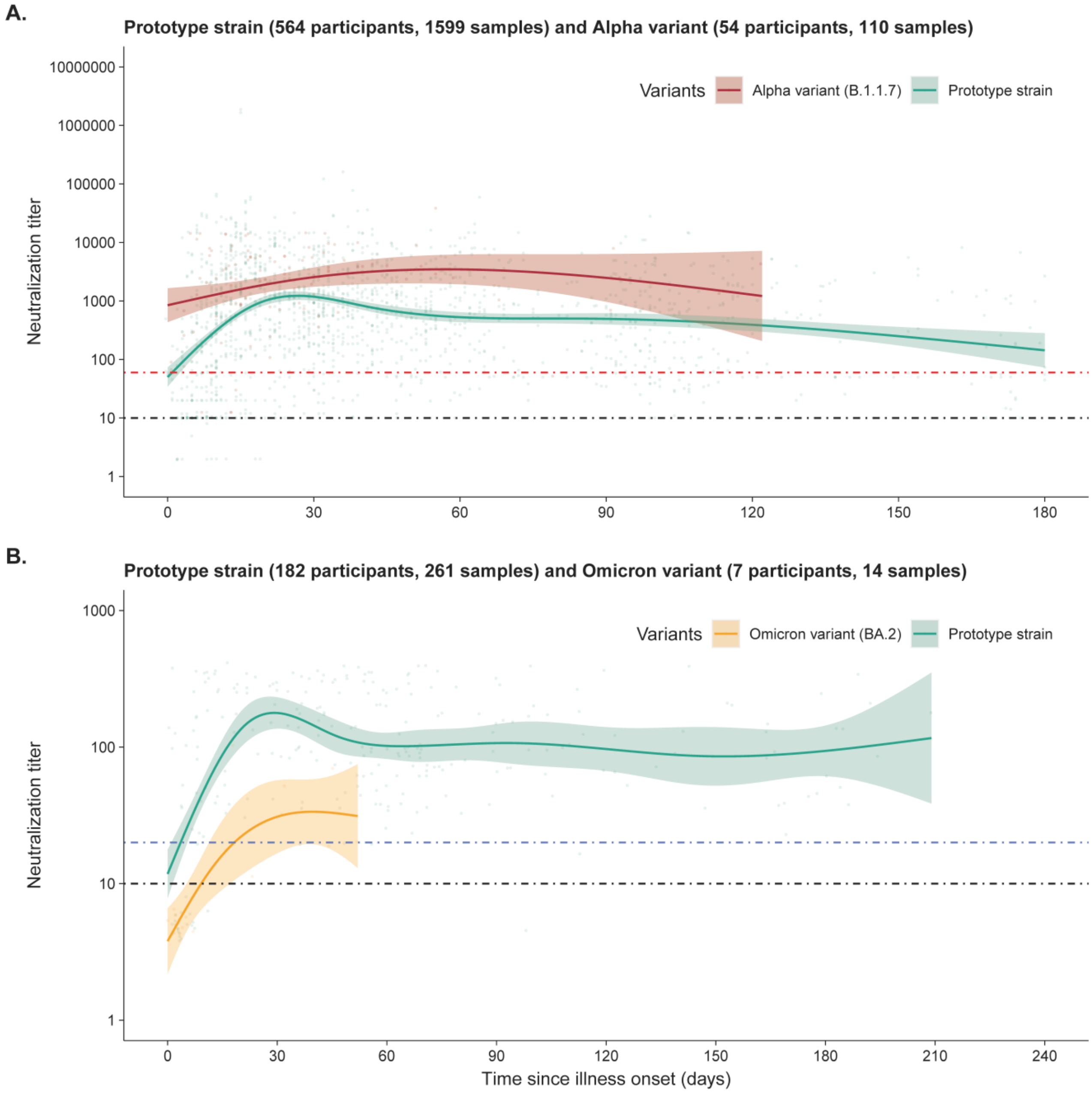

**Figure S4.** **Neutralization antibody responses in prototype strain infections in lentivirus-vector pseudovirus neutralization assays (matched samples)**

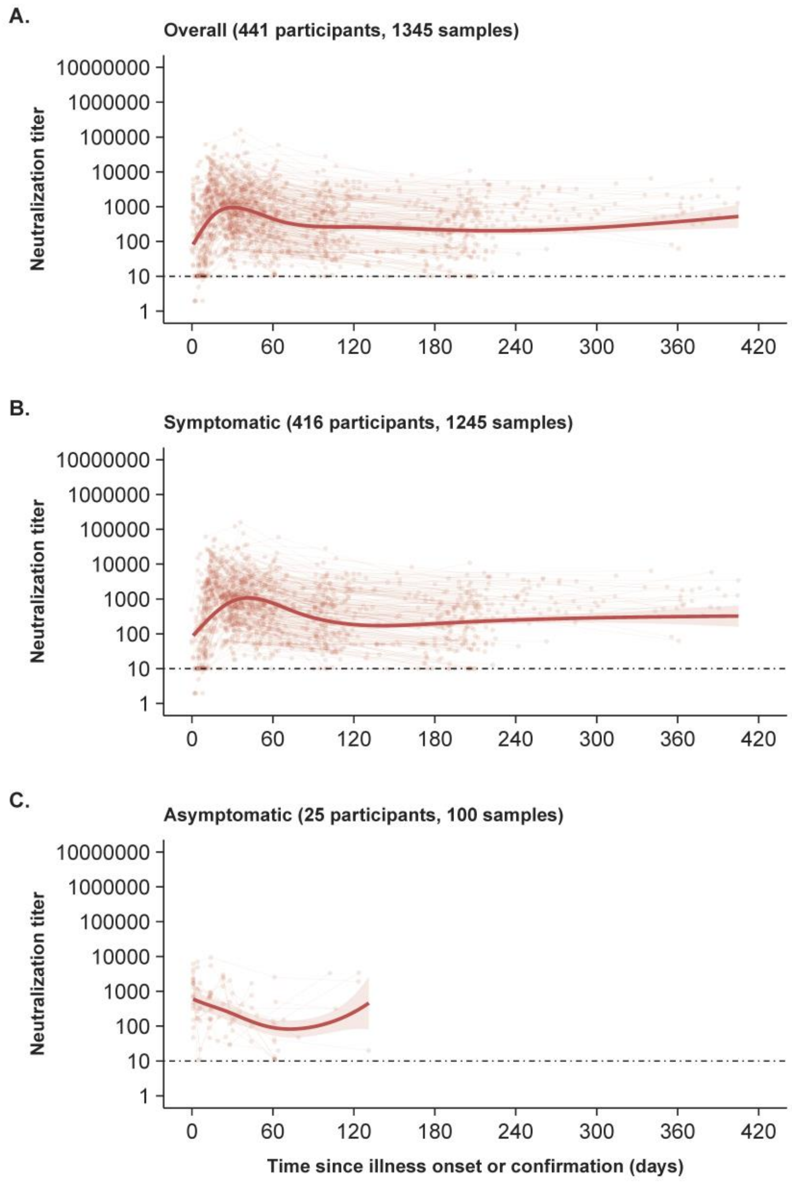

**Figure S5.** **Neutralization antibody responses in prototype strain infections in lentivirus-vector pseudovirus neutralization assays (all samples)**

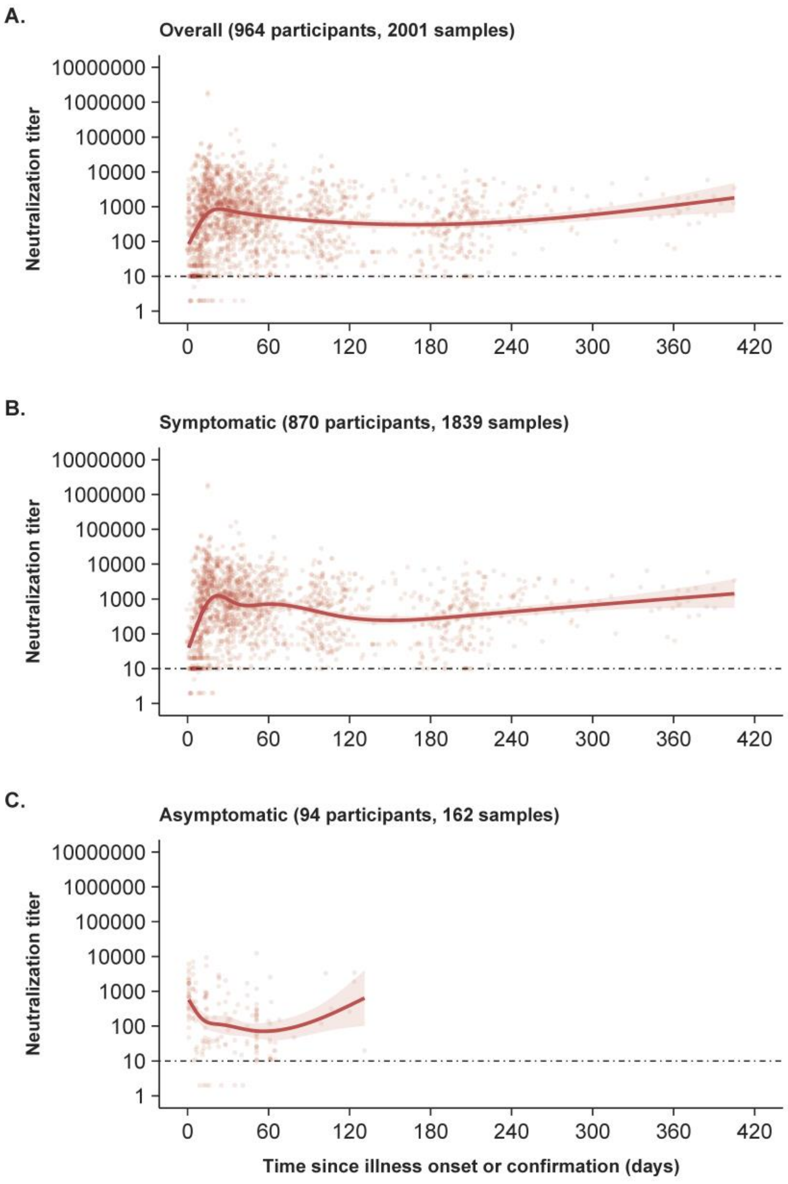

**Figure S6.** **Neutralization antibody responses in prototype strain infections in VSV-vector pseudovirus neutralization assays (all samples)**

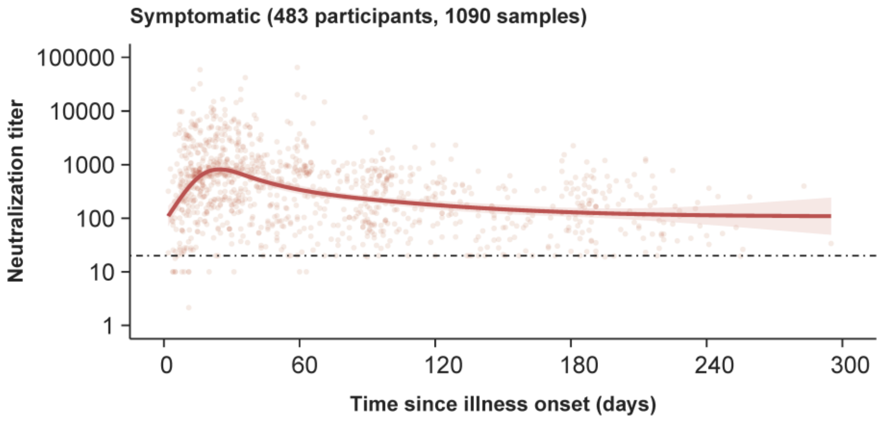
